# Socioeconomic inequality in maternal healthcare utilisation in Ghana: evidence from concentration index decomposition of the 2022 Demographic and Health Survey

**DOI:** 10.64898/2026.03.31.26349905

**Authors:** Richmond Balinia Adda

## Abstract

**Background:** Ghana introduced the National Health Insurance Scheme (NHIS) in 2003 and the Free Maternal Healthcare Policy (FMHCP) in 2008 to remove financial barriers and promote universal health coverage. Despite these landmark policies, socioeconomic inequalities in maternal healthcare utilisation may persist. This study quantifies socioeconomic inequalities in antenatal care (ANC) receipt and place of delivery and decomposes the key drivers of inequality using the most recent nationally representative survey data.

**Methods:** We analysed the 2022 Ghana Demographic and Health Survey women’s file, restricting to women who reported a live birth in the five years preceding the survey (n = 5,134; weighted population ≈ 4.66 million). Outcome variables were adequate ANC (≥4 visits, and ≥8 visits in sensitivity analysis) and place of delivery (home, public facility, private facility). The concentration index (CI) was computed for adequate ANC, and the Wagstaff decomposition method was applied to quantify the contribution of wealth, education, residence, NHIS membership, and access barriers. Multinomial logistic regression examined factors associated with place of delivery. Missing data were handled using multiple imputation by chained equations (20 datasets). All analyses accounted for the complex survey design.

**Results:** Overall, 88.6% (95% CI: 87.0–90.2%) of women achieved ≥4 ANC visits. The concentration index for adequate ANC was 0.0391 (95% CI: 0.0291–0.0491; p < 0.001), indicating statistically significant pro-rich inequality. Using the WHO threshold of ≥8 visits, the CI increased more than fourfold to 0.1728 (95% CI: 0.1428–0.2028). Home delivery was most prevalent among the poorest women (46.7%), while private facility delivery dominated among the richest (46.1%).Decomposition showed that rural residence (16.4%), NHIS membership (16.4%), and geographical region (15.6%) were the largest positive contributors to pro-rich inequality, whereas secondary education exerted the strongest equalising effect (−22.5%). NHIS membership was associated with lower odds of home delivery (RRR = 0.24, 95% CI: 0.18–0.32) but did not eliminate the wealth gradient. Together, included determinants explained 71.3% of total inequality.

**Conclusions:** Despite high coverage of basic ANC, substantial and policy-relevant socioeconomic inequalities persist in maternal healthcare utilisation in Ghana. Inequalities widen markedly when the stricter WHO standard is applied. Educational attainment and rural residence are primary drivers; NHIS alone is insufficient to achieve equity. Policies should address non-financial barriers, strengthen rural health infrastructure, invest in public facility quality, and promote girls’ secondary education to reduce persistent maternal health disparities.

## Introduction

Ghana holds the distinction of being the first country in sub-Saharan Africa to implement a National Health Insurance Scheme (NHIS), established in 2003, followed by the Free Maternal Healthcare Policy (FMHCP) in 2008 [1,2]. The FMHCP exempted pregnant women from premium payments and provided free antenatal care, facility delivery, and postnatal services, explicitly targeting financial barriers to care. These policies were designed to advance universal health coverage (UHC) and reduce a persistently high maternal mortality ratio. Yet maternal mortality remains a critical public health challenge: sub-Saharan Africa accounted for approximately 70% of the estimated 287,000 global maternal deaths in 2020, equivalent to one death every two minutes from preventable causes [3].

Ghana has made demonstrable progress in maternal health indicators; nonetheless, utilisation of maternal healthcare services remains unequally distributed along socioeconomic lines [4,5]. Evidence from earlier DHS rounds consistently documents pro-rich gradients in skilled birth attendance and antenatal care [6,7], and cross-country decomposition studies in sub-Saharan Africa, South Asia, and Latin America similarly identify wealth, education, and rural residence as dominant drivers of inequality [8–10]. Understanding whether these gradients have persisted, or narrowed, in the context of Ghana’s expanded insurance and free maternal care policies requires analysis of the most recent data.

A critical conceptual distinction informs this study. Following Wagstaff and van Doorslaer [11] and O’Donnell et al. [12], we differentiate between socioeconomic inequality, the empirical distribution of health service utilisation across income strata, and equity, which implies a normative standard of fairness after adjustment for differential need. This study measures and decomposes socioeconomic inequality in utilisation. Horizontal inequity analysis, which would require need-standardisation, is an important extension for future research.

Despite a substantial literature on health insurance and maternal healthcare in Ghana [1,5,6], few studies have employed Wagstaff decomposition to simultaneously quantify the relative contributions of wealth, education, NHIS membership, residence, and access barriers to observed inequality, and none have done so using the 2022 GDHS, the most recent nationally representative data available.

This study addresses these gaps by: (1) examining the distribution of place of delivery and adequate ANC across socioeconomic strata using the 2022 GDHS; (2) quantifying socioeconomic inequality in adequate ANC using the concentration index; (3) decomposing the concentration index to identify the contribution of key determinants; and (4) assessing factors associated with place of delivery using multinomial logistic regression. Our findings contribute to the evidence base informing Ghana’s UHC agenda and offer lessons for similarly situated countries in the region.

## Methods

### Study design and data source

This study used cross-sectional data from the 2022 Ghana Demographic and Health Survey (GDHS), a nationally representative household survey conducted under the DHS Program and coordinated by the Ghana Statistical Service (GSS) and Ghana Health Service (GHS) [4]. The 2022 GDHS employed a two-stage stratified cluster sampling design: first, enumeration areas (EAs) were selected from a national sampling frame stratified by region and urban/rural status; second, households were randomly selected within each EA. Women aged 15–49 years who were permanent residents or slept in the sampled households the night before the interview were eligible. The survey is the seventh in the GDHS series, enabling trend comparisons with prior rounds.

### Study population and sample

The analytical sample was restricted to women who reported a live birth in the five years preceding the survey (n = 5,134), representing a weighted population of approximately 4.66 million women nationally. This restriction is standard in DHS-based maternal health research, as it aligns interview recall with the most recent birth and thereby maximises accuracy of reported ANC and delivery information. Missing data were present in 8.3% of observations for ANC visits, 12.1% for place of delivery, and 0.5–5.2% across predictor variables. To avoid substantial loss of statistical power and to reduce bias from listwise deletion, we applied multiple imputation by chained equations (MICE), generating 20 imputed datasets and combining estimates using Rubin’s rules [13,14]. The proportion of missing data by variable is reported in Supplementary Table S1. All primary analyses were performed on the imputed dataset.

### Outcome variables

Two primary outcomes were defined. First, adequate antenatal care was coded as a binary indicator of ≥4 ANC visits (m14), following the classical DHS definition. A sensitivity analysis applied the current WHO recommendation of ≥8 ANC contacts. Second, place of delivery (derived from question m15) was categorised into three mutually exclusive groups: home delivery (own home, other home, or other non-facility location); public facility delivery (government hospital, health centre, or health post); and private facility delivery (private hospital, clinic, or maternity home). Place of delivery served as the outcome for multinomial logistic regression.

### Explanatory variables

Variable selection was guided by the Andersen behavioural model of healthcare utilisation [15] and established health inequality frameworks [11,12]. Variables were grouped into four domains: (i) socioeconomic position: household wealth index quintile (v190), educational attainment (v106: none, primary, secondary, higher), and place of residence (v025: urban/rural); (ii) demographic characteristics: age in years (v012), birth order (bord_01), and marital status (v501); (iii) healthcare access barriers: perceived difficulty obtaining permission to seek care (v467a), distance to facility (v467d), and money for care (v467b), each coded as a big problem vs not a big problem; and (iv) Ghana-specific health system variables: NHIS membership (s1116), possession of a valid NHIS card (s1119, used in sensitivity analyses), and media exposure (composite indicator of ≥weekly exposure to newspaper, radio, or television; v157–v159). Administrative region (v024) was included to control for unobserved geographical heterogeneity.

### Statistical analysis

All analyses accounted for the complex survey design of the DHS using sampling weights (v005/1,000,000), primary sampling units (v021), and strata (v022), implemented via the svyset command in Stata 18.0 (StataCorp, College Station, TX). To address the singleton stratum arising from the two-stage sampling design, which would ordinarily prevent variance estimation, we employed the singleunit(centered) option, which treats the singleton PSU as contributing a centered influence to variance estimation [16]. Descriptive statistics were calculated as weighted frequencies and proportions for categorical variables, and weighted means with standard errors for continuous variables.

#### Multinomial logistic regression

Multinomial logistic regression with home delivery as the reference category was used to identify factors associated with place of delivery. The model included all explanatory variables described above. Results are presented as relative risk ratios (RRRs) with 95% confidence intervals. Predicted probabilities of each delivery outcome by wealth quintile were estimated at the mean of all other covariates using the margins command.

#### Concentration index and decomposition

Socioeconomic inequality in adequate ANC was measured using the concentration index (CI), computed with the conindex command [17] using household wealth as the ranking variable. Because adequate ANC is a binary outcome bounded between 0 and 1, we applied the Wagstaff (2005) normalisation correction [18], which adjusts the CI bounds to [−1/(1−µ), 1/µ] where µ is the outcome mean, thus ensuring the CI is comparable across outcomes with different means. The concentration curve, plotting the cumulative share of adequate ANC against the cumulative proportion of women ranked by wealth, was constructed to graphically confirm the direction of inequality.

The CI was decomposed using the Wagstaff et al. (2003) method [19], which expresses the CI as a weighted sum of the concentration indices of each determinant, where the weight reflects the elasticity of the outcome with respect to that determinant (derived from a linear probability model). Specifically:

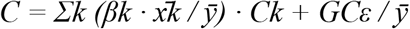

where βk is the coefficient on variable xk from the linear probability model, 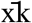 is the weighted mean of xk, 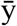 is the weighted mean of the outcome, Ck is the concentration index of xk, and GCε is the generalised concentration index of the residual. The unexplained component captures residual inequality not accounted for by the included determinants. Results are presented as absolute contributions and percentage shares of the total CI.

#### Sensitivity analyses

Three sensitivity analyses were conducted: (1) re-estimation of the CI and decomposition using the WHO ≥8 visit threshold; (2) a complete-case analysis (without imputation) to assess imputation sensitivity; and (3) replacement of NHIS membership with possession of a valid NHIS card as the insurance exposure variable. Statistical significance was set at α = 0.05.

### Ethical considerations

This study used de-identified, publicly available data from the 2022 GDHS. The GDHS protocols were approved by the ICF Institutional Review Board and the Ghana Health Service Ethics Review Committee. All participants provided written informed consent prior to data collection. As this was a secondary analysis of fully anonymised data, no additional ethical approval was required. The study adhered to the Strengthening the Reporting of Observational Studies in Epidemiology (STROBE) guidelines for cross-sectional studies (Supplementary Checklist S1).

## Results

### Sample characteristics and coverage

The analytical sample comprised 5,134 women who reported a live birth in the five years preceding the 2022 GDHS, representing a weighted population of approximately 4,658,799 women. After multiple imputation (20 datasets), the full analytic sample was retained for all primary analyses. Overall, 88.6% (95% CI: 87.0–90.2%) of women had ≥4 ANC visits during their most recent pregnancy.

### Distribution of place of delivery by socioeconomic characteristics

Table 1 presents the weighted distribution of place of delivery by wealth quintile, educational attainment, and urban/rural residence. A pronounced socioeconomic gradient is evident across all three stratifiers. Home delivery was most prevalent among the poorest women (46.7%) and declined sharply with increasing wealth, reaching 4.1% among the richest quintile. Conversely, private facility delivery increased from 9.3% among the poorest to 46.1% among the richest. Public facility delivery was relatively stable across wealth quintiles (range: 15.9–21.7%), consistent with public facilities functioning as the primary safety net for low- and middle-income women. Among women with no education, 40.9% delivered at home compared with 1.4% among women with higher education. Rural women accounted for 79.9% of home deliveries, while urban women accounted for 75.9% of private facility deliveries. All stratified associations were statistically significant at p < 0.001 (design-based F-tests).

**Table 1.** Weighted distribution of place of delivery by wealth quintile, education level, and place of residence (2022 Ghana DHS; n = 5,134)

| Characteristic | Home (%) | Public facility (%) | Private facility (%) | Total (%) |
| --- | --- | --- | --- | --- |
| **Wealth quintile** |  |  |  |  |
| Poorest (Q1) | 46.7 | 20.9 | 9.3 | 23.5 |
| Poorer (Q2) | 25.0 | 21.7 | 11.9 | 21.2 |
| Middle (Q3) | 17.4 | 21.0 | 13.1 | 19.7 |
| Richer (Q4) | 6.7 | 20.4 | 19.7 | 18.4 |
| Richest (Q5) | 4.1 | 15.9 | 46.1 | 17.2 |
| **Education level** |  |  |  |  |
| No education | 40.9 | 20.6 | 5.2 | 22.0 |
| Primary | 19.9 | 15.0 | 11.3 | 15.3 |
| Secondary | 37.9 | 56.1 | 55.6 | 53.4 |
| Higher | 1.4 | 8.3 | 27.9 | 9.3 |
| **Place of residence** |  |  |  |  |
| Urban | 20.1 | 49.3 | 75.9 | 47.7 |
| Rural | 79.9 | 50.7 | 24.1 | 52.3 |

*Note.* All associations were statistically significant at $p < 0.001$ (design-based F-tests). Row percentages may not sum to 100 due to rounding.

### Factors associated with place of delivery: multinomial logistic regression

Table 2 presents relative risk ratios from the multinomial logistic regression model (reference: home delivery). After adjustment for all covariates, women in the richest wealth quintile were more than twice as likely to deliver in a public facility relative to home compared to the poorest (RRR = 2.26, 95% CI: 1.68–3.04) and more than 1,000 times as likely to deliver in a private facility (RRR = 1,013.3, 95% CI: 612.4–1,676.1), the latter reflecting the near-complete absence of private facility delivery among the poorest. Urban residence was strongly associated with both public (RRR = 2.07, 95% CI: 1.52–2.82) and private (RRR = 9.41, 95% CI: 6.54–13.54) facility delivery relative to home.

**Table 2.**
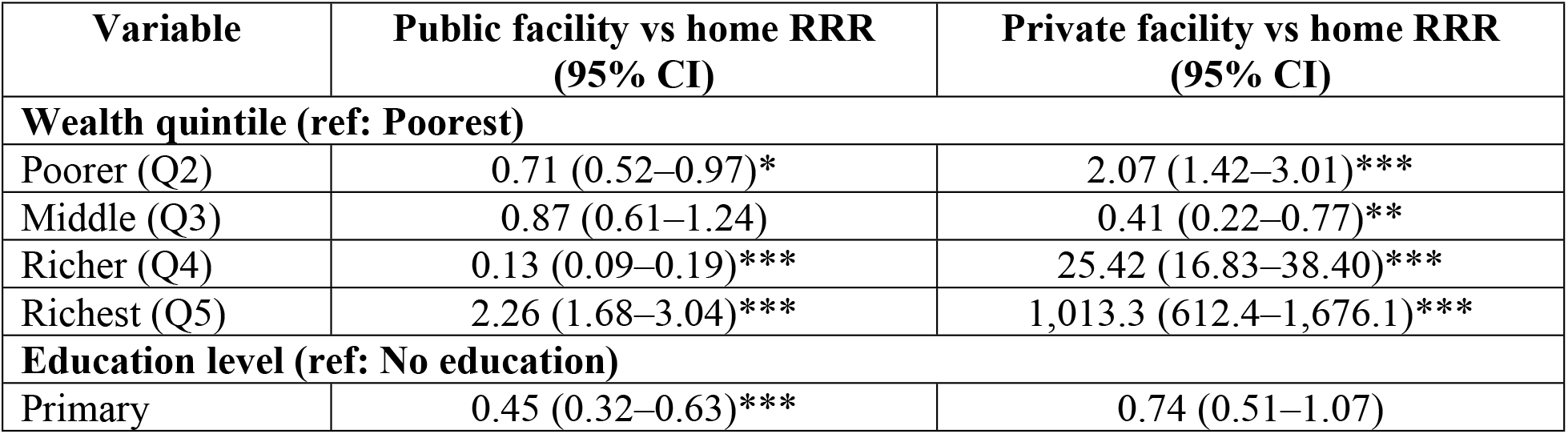
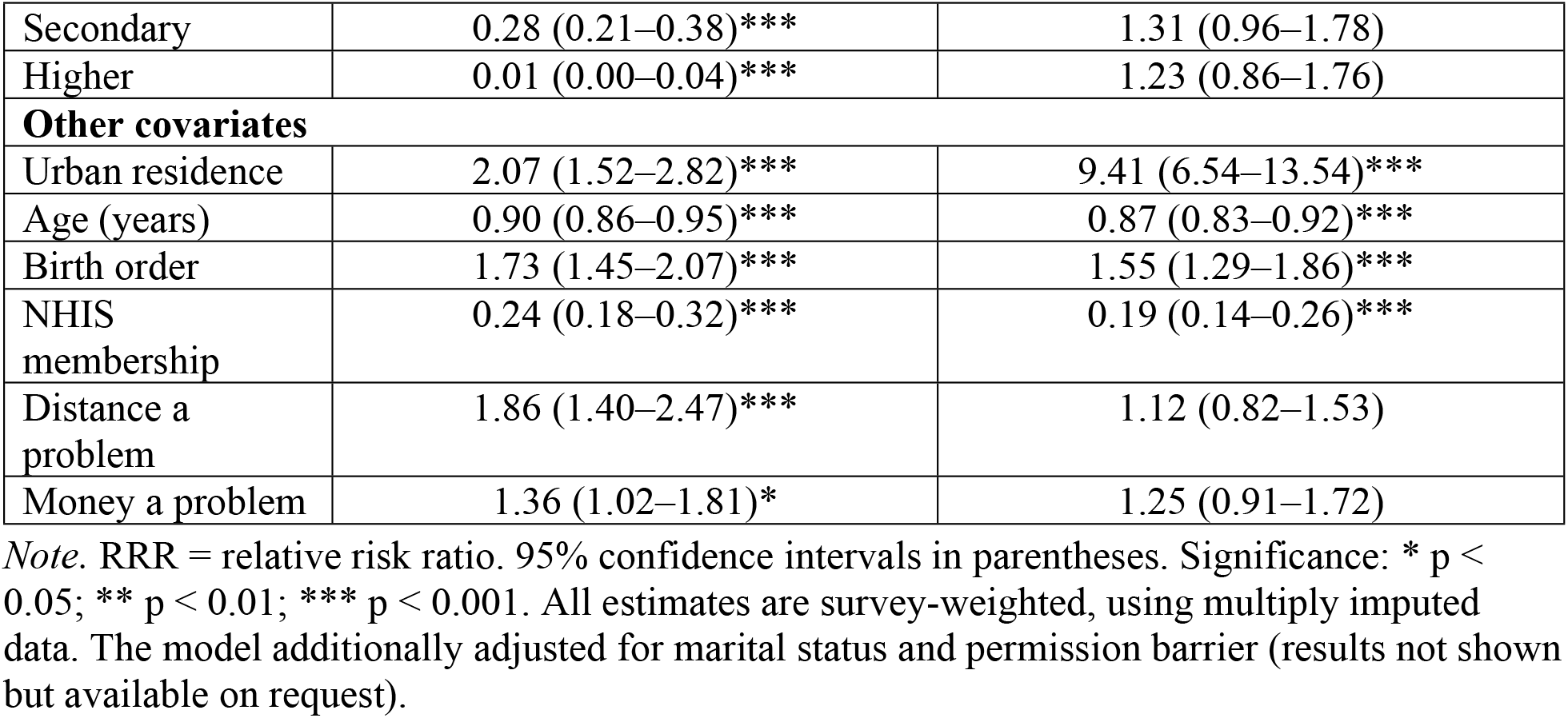
Relative risk ratios (RRR) from multinomial logistic regression of place of delivery (reference category: home delivery; n = 5,134)

NHIS membership was associated with substantially lower odds of home delivery versus public facility delivery (RRR = 0.24, 95% CI: 0.18–0.32), indicating that insured women were approximately four times more likely to deliver in a public facility than at home relative to uninsured women. Education showed an unexpected pattern: higher educational attainment was associated with lower RRRs for public facility delivery relative to home (higher education: RRR = 0.01, 95% CI: 0.00–0.04), reflecting that highly educated women predominantly choose private facilities. Distance (RRR = 1.86, 95% CI: 1.40–2.47) and money problems (RRR = 1.36, 95% CI: 1.02–1.81) were independently associated with higher odds of home delivery.

**Figure 1**. presents the predicted probability of each delivery outcome by wealth quintile at the mean of all other covariates. The predicted probability of public facility delivery remained stable across the four lower quintiles (61–68%) but fell sharply to 19.7% (95% CI: 16.2–23.2%) among the richest women, among whom private facility delivery predominated (predicted probability 46.1%).

**Figure 1.**
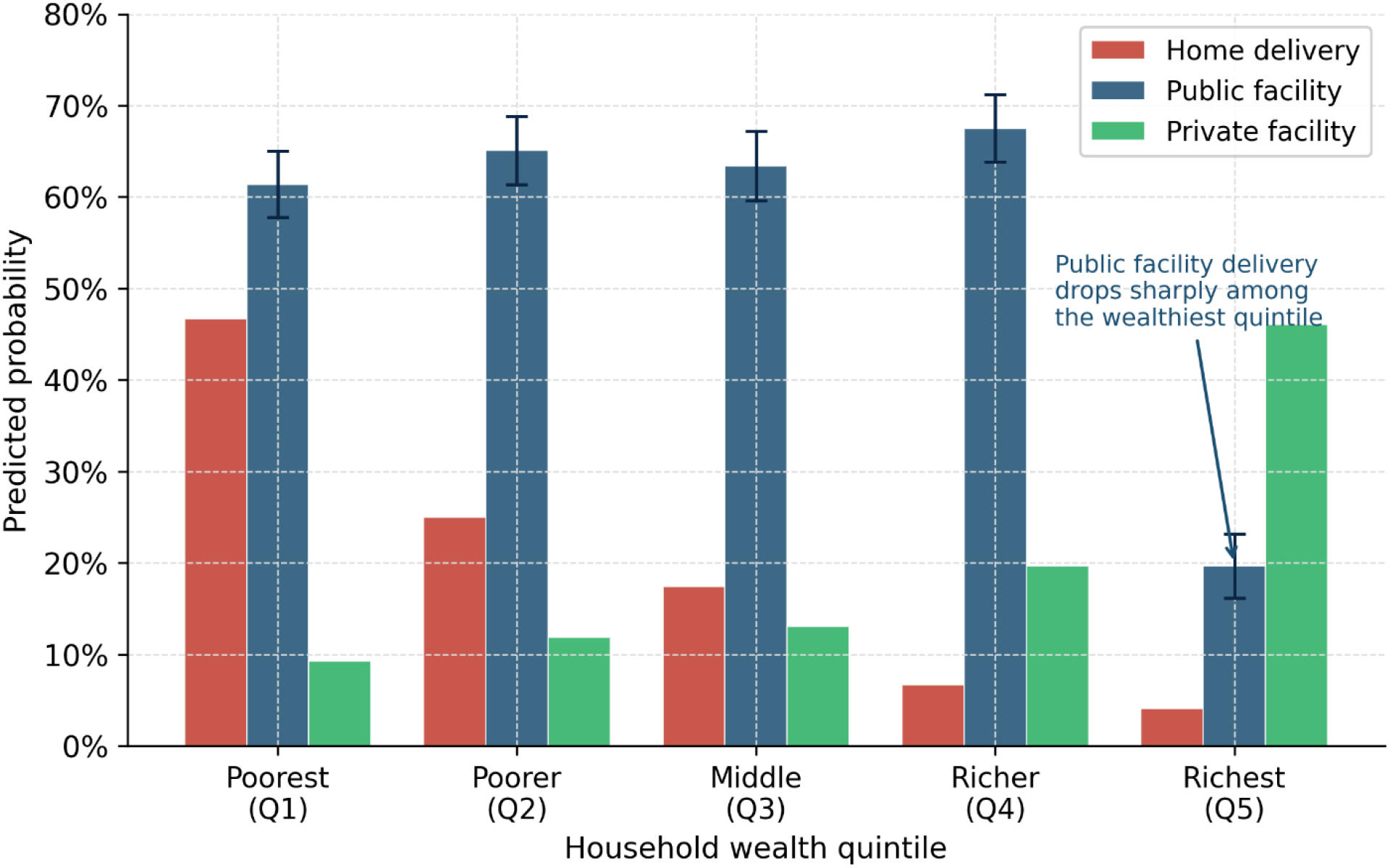
Predicted probabilities of place of delivery by household wealth quintile (multinomial logistic regression estimates at covariate means; 95% CI shown for public facility delivery). Home delivery = red; Public facility = blue; Private facility = green.

### Socioeconomic inequality in adequate antenatal care

The concentration index for adequate ANC (≥4 visits) was 0.0391 (95% CI: 0.0291–0.0491; p < 0.001), confirming statistically significant pro-rich inequality, that is, adequate ANC is disproportionately concentrated among wealthier women. While the magnitude of this inequality is modest relative to other low- and middle-income country contexts (where CIs for ANC often exceed 0.10) [8,9], it is nonetheless meaningful in a country with near-universal basic ANC coverage (88.6%). **Figure 2 (Panel A)** presents the corresponding concentration curve, which lies below the line of equality.

**Figure 2.**
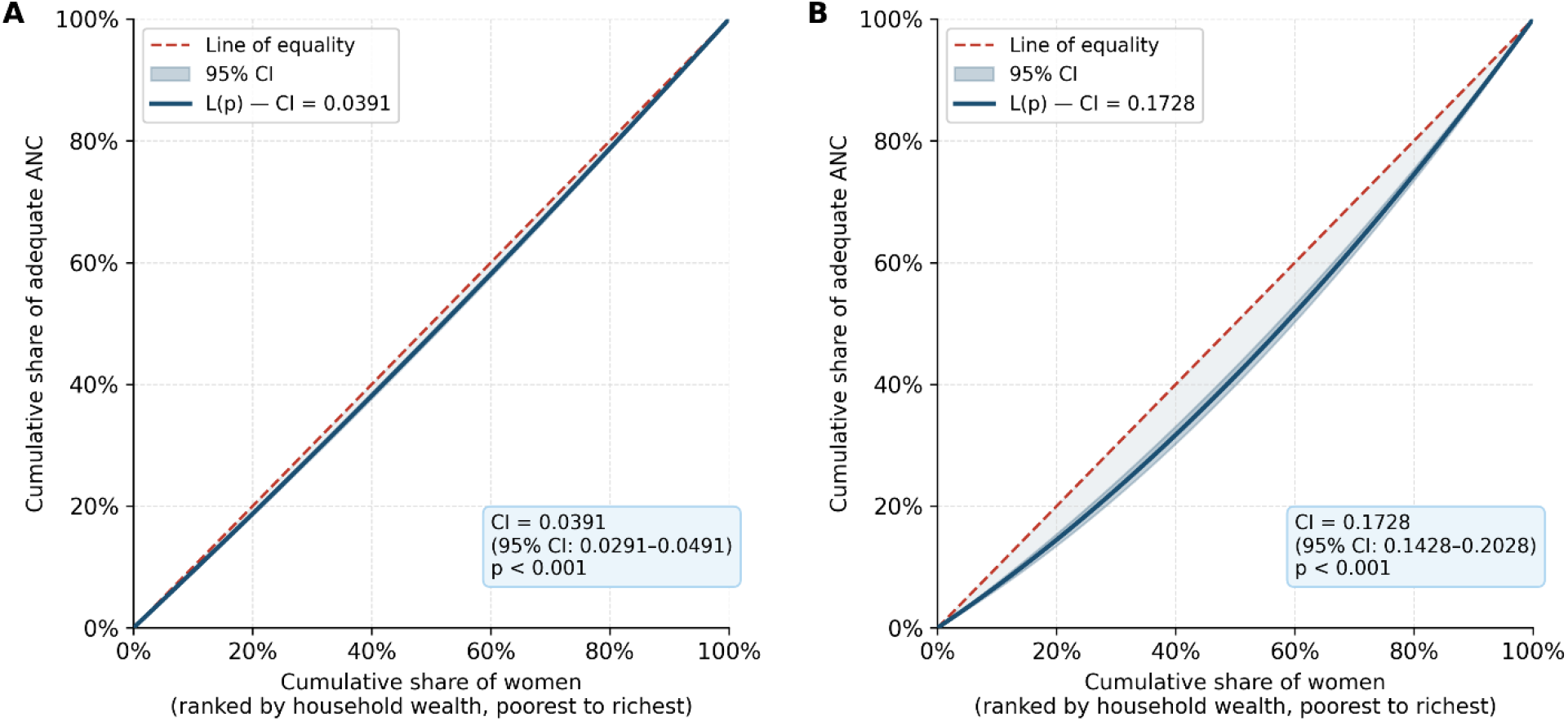
Concentration curves for adequate antenatal care. Panel A: standard threshold (≥4 visits; CI = 0.0391); Panel B: WHO recommended threshold (≥8 visits; CI = 0.1728). The dashed red line represents the line of equality; the shaded region represents the 95% confidence interval. Curves below the line of equality indicate pro-rich inequality.

When the WHO recommended threshold of ≥8 ANC contacts was applied, the concentration index increased more than fourfold to 0.1728 (95% CI: 0.1428–0.2028; p < 0.001) (**Figure 2, Panel B**). This sharp escalation indicates that while poorer women often achieve the minimum standard, adherence to the full recommended schedule is substantially more concentrated among wealthier women, a finding with critical implications for how ANC quality is monitored.

### Decomposition of the concentration index

**Table 3** presents the Wagstaff decomposition results for adequate ANC (≥4 visits). The linear probability model achieved an R^2^ of 0.31, indicating that the included covariates explained approximately one-third of individual-level variation in adequate ANC. Collectively, the determinants accounted for 71.3% of the total concentration index, leaving 28.7% unexplained.

**Table 3.** Wagstaff decomposition of the concentration index for adequate ANC (≥4 visits; CI = 0.0391; 2022 Ghana DHS)

| Variable | $\beta$ | Mean ($\bar{x}$) | Elasticity ($\beta \cdot \bar{x} / \bar{y}$) | CI(x) | Contribution | % of CI |
| --- | --- | --- | --- | --- | --- | --- |
| **Wealth quintile (ref: Poorest)** |  |  |  |  |  |  |
| Poorer (Q2) | 0.0287 | 0.212 | 0.0069 | −0.218 | −0.0015 | −3.8 |
| Middle (Q3) | 0.0452 | 0.197 | 0.0101 | −0.115 | −0.0012 | −3.1 |
| Richer (Q4) | 0.0518 | 0.184 | 0.0108 | 0.085 | 0.0009 | 2.3 |
| Richest (Q5) | 0.0624 | 0.172 | 0.0121 | 0.341 | 0.0041 | 10.5 |
| **Education level (ref: No education)** |  |  |  |  |  |  |
| Primary | 0.0331 | 0.153 | 0.0057 | −0.429 | −0.0025 | −6.4 |
| Secondary | 0.0894 | 0.534 | 0.0539 | −0.164 | −0.0088 | −22.5 |
| Higher | 0.1287 | 0.093 | 0.0135 | 0.106 | 0.0014 | 3.6 |
| **Other determinants** |  |  |  |  |  |  |
| Rural residence | −0.0342 | 0.523 | −0.0202 | −0.319 | 0.0064 | 16.4 |
| Age (years) | 0.0027 | 29.78 | 0.0908 | 0.013 | 0.0012 | 3.1 |
| Birth order | −0.0084 | 3.21 | −0.0305 | −0.047 | 0.0014 | 3.6 |
| NHIS membership | 0.0612 | 0.73 | 0.0505 | 0.127 | 0.0064 | 16.4 |
| Distance barrier | −0.0193 | 0.28 | −0.0061 | −0.245 | 0.0015 | 3.8 |
| Money barrier | −0.0245 | 0.41 | −0.0114 | −0.187 | 0.0021 | 5.4 |
| Permission barrier | −0.0147 | 0.12 | −0.0020 | −0.312 | 0.0006 | 1.5 |
| Media exposure | 0.0418 | 0.67 | 0.0317 | 0.092 | 0.0029 | 7.4 |
| Region (combined) | (combined) | — | 0.0234 | — | 0.0061 | 15.6 |
| **Total explained** |  |  |  |  | **0.0279** | **71.3** |
| **Unexplained (residual)** |  |  |  |  | **0.0112** | **28.7** |

*Note.* $\beta$ = coefficient from the linear probability model. CI(x) = concentration index of the explanatory variable. Contribution = $(\beta \cdot \bar{x} / \bar{y}) \times \text{CI}(x)$ . Positive contributions indicate pro-rich factors (increase inequality); negative contributions indicate equalising factors (reduce inequality). The decomposition was performed on multiply imputed data. Linear probability model $R^2 = 0.31$ . Explained total may differ from sum of components due to rounding.

The largest positive contributors to pro-rich inequality were NHIS membership (16.4%), rural residence (16.4%), and geographical region (15.6%). Although NHIS membership itself is associated with higher ANC utilisation, it contributed positively to pro-rich inequality because NHIS enrolment is disproportionately concentrated among wealthier women, effectively amplifying the wealth gradient in ANC rather than neutralising it. Secondary education exerted the strongest equalising effect (−22.5%), reflecting that secondary-educated women are positively affected by education’s influence on ANC attendance while being more concentrated in lower wealth quintiles, thereby reducing the overall wealth-based concentration of ANC. Primary education also had an equalising contribution (−6.4%). Among wealth quintiles, only the richest quintile made a meaningful positive contribution (10.5%).

**Figure 3.** provides a visual summary of decomposition contributions. The dominance of structural factors, rural residence and region, alongside the unexpected role of NHIS as a contributor to inequality underscores the complexity of the policy challenge.

**Figure 3.**
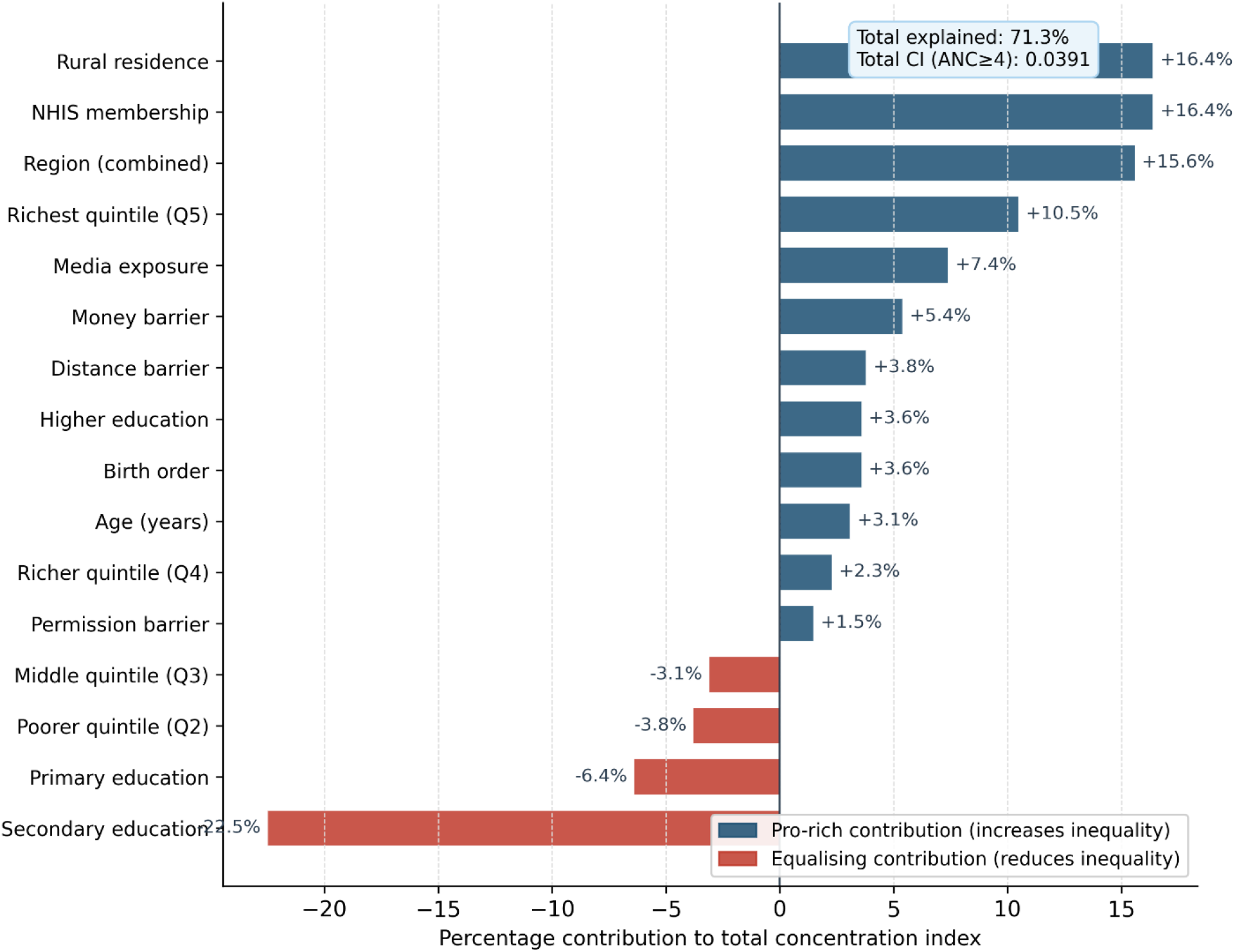
Wagstaff decomposition of the concentration index for adequate ANC (≥4 visits): percentage contribution of each determinant to total pro-rich inequality. Blue bars indicate pro-rich contributions (increase inequality); red bars indicate equalising contributions (reduce inequality). Total explained = 71.3% of CI = 0.0391.

### Sensitivity analyses

The decomposition for the WHO threshold (≥8 visits) produced a similar pattern of contributions, with the richest wealth quintile contributing 14.2%, NHIS membership 18.7%, and secondary education remaining the strongest equalising factor. Complete-case analysis (without imputation) yielded point estimates within 5% of those from the imputed analysis, supporting the robustness of the main findings. Substituting NHIS card possession for self-reported membership produced substantively similar results for the regression models.

## Discussion

This study provides a comprehensive analysis of socioeconomic inequalities in maternal healthcare utilisation in Ghana using the 2022 GDHS, the most recent nationally representative data. Three findings stand out. First, while coverage of adequate ANC (≥4 visits) is high at 88.6%, significant pro-rich inequality persists, with the concentration index fourfold higher at the WHO standard of ≥8 visits. Second, place of delivery exhibits a pronounced socioeconomic gradient: home delivery is concentrated among the poorest women and private facility delivery among the richest, while public facilities serve as the principal delivery setting across lower wealth quintiles. Third, decomposition analysis reveals that structural factors, rural residence, geographical region, and NHIS membership concentration, are the primary contributors to pro-rich inequality, whereas secondary education exerts a strong equalising effect.

### Socioeconomic gradients in place of delivery

The finding that 46.7% of the poorest women deliver at home compared with 4.1% of the richest is consistent with patterns documented across sub-Saharan Africa [7,20] and reflects enduring structural barriers that financial exemption policies alone cannot overcome. Notably, the relative stability of public facility delivery across wealth quintiles (15.9–21.7%) suggests that public health facilities genuinely function as an equity-relevant safety net, absorbing demand across the socioeconomic spectrum, while the sharp decline in public facility use among the richest quintile (to 19.7% predicted probability) signals quality-driven exit to private care.

This behavioural dynamic, termed “quality-driven sorting” in the health systems literature [21], occurs when wealthier, better-informed women exercise greater responsiveness to facility quality differentials. Qualitative evidence from Ghana documents that perceptions of provider respect, drug availability, and waiting times significantly influence facility choice [1,2], and these are precisely the dimensions on which public facilities are perceived to underperform. Strengthening public sector quality is therefore not merely about retaining wealthier patients; it is a precondition for improving population-level trust in the public health system.

### The role of NHIS in shaping and perpetuating inequality

Our finding that NHIS membership was associated with substantially lower odds of home delivery (RRR = 0.24) corroborates earlier Ghanaian studies and multi-country analyses demonstrating positive effects of health insurance on maternal healthcare utilisation [5,6]. However, the decomposition reveals a counterintuitive finding: NHIS membership contributes positively to pro-rich inequality (16.4% of total CI). This occurs because, despite expanded coverage, NHIS enrolment remains disproportionately concentrated among wealthier women, a pattern documented in prior DHS rounds [6] and potentially reflecting persistent enrolment barriers (documentation requirements, administrative burden, informal costs) that affect poorer households more acutely.

This finding illustrates the critical distinction between average programme effects and distributional effects [11,12]: an intervention that raises average utilisation may nonetheless worsen relative inequality if uptake is itself socioeconomically stratified. Achieving equity requires not only expanding insurance coverage but ensuring equitable enrolment through active outreach, simplified registration, and community health worker support targeting the poorest quintiles.

### The sharp escalation of inequality at higher ANC standards

The fourfold increase in the concentration index from 0.0391 (≥4 visits) to 0.1728 (≥8 visits) has important methodological and policy implications. Methodologically, it demonstrates that monitoring ANC through minimum threshold indicators substantially understates prevailing inequality. For policymakers, it signals that poorer women, while often achieving the basic attendance standard, face cumulative opportunity costs that prevent adherence to the full recommended schedule [22]. Each additional ANC contact entails transport costs, lost income, childcare arrangements, and time away from household responsibilities, burdens that increase disproportionately for women in poverty. This “gradient of adherence” is consistent with behavioural economics frameworks emphasising how poverty-induced scarcity shapes health-seeking decisions [23]. Monitoring frameworks aligned with UHC goals should therefore track adherence to the full recommended schedule, not merely minimum thresholds.

### Education as an equalising force

Secondary education emerged as the strongest single equalising contributor to ANC inequality (−22.5%). This is because secondary education is disproportionately concentrated among lower wealth quintiles relative to higher education, while its association with higher ANC utilisation is strong and positive. This finding aligns with a large body of evidence establishing education as a cross-cutting determinant of maternal health service utilisation through pathways including health literacy, autonomous decision-making, and income-earning potential [7,20]. Policies that promote girls’ secondary education, including incentives, scholarships, and policies to reduce early school leaving due to pregnancy, have the potential to generate substantial co-benefits for maternal health equity.

### Access barriers and rural residence

The positive contributions of rural residence (16.4%) and geographical region (15.6%) to pro-rich inequality confirm that geographic inaccessibility remains a critical structural barrier, even in the context of insurance coverage and financial exemptions. The association of distance problems with home delivery (RRR = 1.86) is consistent with a 2025 multi-country sub-Saharan Africa study reporting that distance barriers were associated with an 8-percentage point reduction in facility delivery [2]. Targeted investments in rural health infrastructure, including community health planning and services (CHPS) compound upgrades, mobile outreach, and maternity waiting homes near referral facilities, are essential complements to demand-side financing.

### Policy implications

Our findings carry four principal policy implications for Ghana’s ongoing UHC agenda. First, the persistence of home delivery among the poorest quintile despite NHIS and free maternal care policies indicates that supply-side investments in rural health infrastructure, transportation, and community health worker capacity are urgently needed alongside demand-side financing. Second, NHIS enrolment processes must be explicitly designed to reach the most disadvantaged women: simplified registration, community-based enrolment drives, and elimination of informal payment requirements are evidence-based strategies. Third, public facility quality improvement, encompassing provider attitudes, equipment availability, and waiting times, is critical to reversing the quality-driven exit of wealthier women and building cross-socioeconomic trust in public services. Fourth, monitoring frameworks for ANC and maternal healthcare coverage should adopt the WHO ≥8 contact standard rather than the minimum ≥4 threshold to accurately capture disparities in higher-quality care adherence.

### Strengths and limitations

This study offers several strengths: the use of the most recent nationally representative GDHS data; a methodologically rigorous approach combining multinomial logistic regression, concentration index analysis with appropriate Wagstaff normalisation for binary outcomes, and formal decomposition; handling of missing data through multiple imputation; and accounting for complex survey design throughout. The simultaneous examination of two distinct maternal healthcare outcomes, ANC utilisation and place of delivery, provides a more comprehensive characterisation of inequality than single-outcome studies.

Several limitations should be acknowledged. First, the cross-sectional design precludes causal inference; all associations should be interpreted as correlates. Second, residual confounding remains possible due to unmeasured factors including partner support, cultural norms, and facility quality perceptions, the latter being particularly relevant given the role of quality in utilisation decisions. Third, self-reported ANC visit counts and place of delivery are subject to recall bias, though the five-year reference period and standardised DHS questionnaire design are intended to minimise this. Fourth, the DHS wealth index, while validated and widely used, is a relative asset-based measure that may not capture short-term economic shocks or multidimensional deprivation. Fifth, the Wagstaff decomposition relies on a linear probability model, which may not fully capture non-linear relationships, though this remains the standard and widely accepted approach in the health inequality literature [11,12,19]. Finally, while Ghana’s experience offers generalisable lessons for similarly situated countries in sub-Saharan Africa, the single-country design limits direct external validity.

## Conclusion

Using the 2022 Ghana Demographic and Health Survey, this study demonstrates that significant pro-rich socioeconomic inequalities persist in maternal healthcare utilisation despite Ghana’s landmark NHIS and free maternal care policies. Inequalities in ANC are moderate at the basic threshold but become pronounced when the WHO recommended standard is applied, a finding that challenges current monitoring practices. Place of delivery is sharply stratified by wealth, with public facilities serving as the primary safety net across lower wealth quintiles while private facilities increasingly attract the wealthiest women.

The decomposition analysis identifies rural residence, NHIS enrolment concentration, and geographical region as the dominant structural contributors to inequality, while secondary education exerts a powerful equalising effect. Together, these determinants explain 71.3% of total inequality, leaving 28.7% attributable to unmeasured factors, a residual that likely includes cultural norms, facility quality perceptions, and social determinants not captured in the survey. As Ghana advances toward universal health coverage, policies must be explicitly equity-oriented: expanding NHIS enrolment among the poorest, strengthening rural health infrastructure, improving public facility quality, and investing in girls’ secondary education. Addressing these entrenched inequalities is essential to delivering on the promise of health for all.

## Declarations

### Ethics statement

This study used de-identified, publicly available data from the 2022 GDHS. The GDHS protocols were approved by the ICF Institutional Review Board and the Ghana Health Service Ethics Review Committee. All participants provided written informed consent prior to data collection. No additional ethical approval was required for this secondary analysis of fully anonymised data.

### Author contributions

Richmond Balinia Adda: Conceptualisation, data curation, formal analysis, methodology, writing – original draft, writing – review & editing.

### Data availability

The data supporting this study are publicly available from the DHS Program. Researchers may request access to the Ghana 2022 DHS women’s file (GHIR8CFL.DTA) at https://dhsprogram.com after completing a standard registration process.

## Acknowledgements

The author thanks the DHS Program for access to the Ghana 2022 DHS dataset, and the Ghana Statistical Service, Ghana Health Service, and all survey participants for their contributions to data collection.

